# Early Improvement of Acute Respiratory Distress Syndrome in Patients with COVID-19: Insights from the Data of ICU Patients in Chongqing, China

**DOI:** 10.1101/2020.07.15.20154047

**Authors:** Zhu Zhan, Xin Yang, Hu Du, Chuanlai Zhang, Yuyan Song, Xiaoyun Ran, An Zhang, Mei Yang

## Abstract

Acute respiratory distress syndrome (ARDS) may be the main cause of death in patients with coronavirus disease 2019 (COVID-19). Herein, we retrospect clinical features, outcomes and ARDS characteristics of 75 intensive care unit (ICU) patients with COVID-19 in Chongqing, China. We found a 5.3% case fatality rate of the ICU patients in Chongqing. 93% patients developed ARDS during the intensive care, and more than half were moderate. However, most of the patients (55%) supported with high flow nasal cannula (HFNC) oxygen therapy, but not mechanical ventilation. Nearly one third of patients with ARDS got an early improvement (eiARDS), and the rate is much higher than the other causes of ARDS in a previous study. Patients with eiARDS had a higher survival rate and lower length of ICU stay. The age (< 55 years) is an independent predictor for the eiARDS, and stratification of COVID-19 patients by age is recommended.

## Introduction

On December 2019, Wuhan (Hubei province, China) reported a cluster cases of unknow cause of pneumonia, which were later identified as coronavirus disease 2019 (COVID-19)(1). This contagious disease was caused by severe acute respiratory syndrome coronavirus 2 (SARS-CoV-2), and was declared to be a worldwide pandemic by World Health Organization (WHO) on March11, 2020(2). As of 23 May 2020, a total of more than 5 000 000 cases and 330 000 deaths had been reported worldwide(3).

The leading cause of COVID-19 death may be the severe respiratory failure caused by acute respiratory distress syndrome (ARDS)(4). Because according to some autopsy results, the lesions are primarily in the lungs, characterized by diffuse alveolar damage. Other organs, such as the heart tissue, have no obvious histological changes(5–7). Previous studies reported that about 48.6% of patients with COVID-19 had ARDS, among which 29% patients had died, and the mortality rate increased with the severity of ARDS(8,9). Therefore, it is needed to fully understand the features of ARDS in patients with COVID-19.

In this study, we described epidemiology, clinical features, laboratory findings, treatments, and outcomes of intensive care unit (ICU) patients with COVID-19 in Chongqing, China, the adjoining areas of Hubei province. And we identified a subphenotype of ARDS – early improvement of ARDS, which occurred in about one third patients, and would predict a favorable clinical outcome.

## Methods

### Study design and participants

This retrospective cohort study included two cohorts of ICU patients from Chongqing public health medical center and Chongqing three gorges central hospital (Chongqing, China), both of which are designated hospital to treat patients with COVID-19 in Chongqing. Patients who admitted to ICU between Jan 21, 2020 (time of the first patient admitted) and March 15, 2020 (time of the last patient discharged in the first wave), were enrolled in our study.

Patients with COVID-19 were confirmed by the positive real-time reverse transcriptase– polymerase chain reaction (RT-PCR) assay for nasal and pharyngeal swab specimens according to the WHO guidance. The severity of COVID-19 was judged according to the Fifth Revised Trial Version of the Novel Coronavirus Pneumonia Diagnosis and Treatment Guidance of China(10). Those who met the any following criteria were defined as severe-type: (1) respiratory resting state, and (3) arterial blood oxygen partial pressure (PaO2) /oxygen concentration (FiO2) ≤ 300 mm Hg. Those who met one of the following criteria were defined as critically ill-type: (1) mechanical ventilation needed for respiratory failure, (2) shock, and (3) intensive care needed owing to other organ failure.

The study was approved by the Research Ethics Commission of the second affiliated hospital of Chongqing medical university, Chongqing public health medical center and Chongqing three gorges central hospital. Written informed consent was waived by the Ethics Commission of the designated hospital for emerging infectious disease.

### Data collection

Both of the two designated hospitals are Grade A hospitals in China, and all the case data can be found in the electronic case system. Epidemiological, demographic, symptoms, underlying diseases, comorbidities, treatments, clinical course and outcome data of the patients were recorded in a spreadsheet. The signs, arterial blood gas analysis, laboratory data, acute physiology and chronic health evaluation II (APACHE II) and sequential organ failure assessment (SOFA) score were collected at the certain time (Day0: admission in hospital, Day1: admission in ICU, Day3, Day7 and Day14) for each patient. If there was no question about the case data, the doctor in charge was promptly inquired. As the data collection was completed, another doctor was responsible for checking and integrating. The proportion of pneumonia volume was calculated according to the pulmonary infection assisted diagnosis system (V1.7.0.1) based on the Computed Tomography image.

### Definition

ARDS was diagnosed according to the Berlin Definition(11). Liver injury was diagnosed according to the following criteria: alanine aminotransferase (ALT) > 3 upperlimit of normal (ULN) or aspartate aminotransferase (AST) > 3ULN or Total bilirubin (TBIL) > 2ULN, regardless of chronic liver disease(12). Acute kidney injury was diagnosed on the basis of serum creatinine(13). Cardiac injury was diagnosed if the serum concentration of hypersensitive cardiac troponin T (hsTNT) was above the upper limit of the reference range (>14 pg/mL). Time of viral shedding was defined as when two consecutive SARS-CoV-2 PCR assays at least 24 h apart were negative.

### Statistical analysis

SPSS26.0 (IBM SPSS Statistics, IBM Corporation) was used as the statistical analysis tool. The continuous variables that met the normal distribution are presented as mean ± standard deviation, and the independent student’s t test was used for comparison between two groups. The continuous variables that do not meet the normal distribution are presented as the median (interquartile ranges, IQR), and the Mann-Whitney U test was used between two groups. Categorical variables are summarized by using frequencies and percentages, and the L2 test or the Fisher exact test was used among two or more groups; We performed bivariate analyses to identify the predictors for early improvement of ARDS, variables with a P value < 0.05 in the univariate analysis were entered into multivariate logistic regression analysis. All tests were two-sided and p < 0.05 were considered statistically significant.

## Result

### Clinical characteristics of ICU patients in Chongqing, China

From 21 January to 15 March 2020, Chongqing reported 576 new cases of COVID-19, with 6 deaths. 75 ICU patients from two hospitals were included in this study, with 48 severe and 27 critically ill patients, and 4 death.

The comparison of clinical characteristics between these two groups is shown in Table 1. The median age of the 75 patients was 57 years (IQR 25-75), and no bias in the sex ratio. Smoking was more prevalent in critically ill patients (30%) than the severe ones (2%, p = 0.002). 16 (21%) patients had a history of exposure to Hubei province, 24 (32%) patients contacted with patients from Hubei, 17 (23%) patients contacted with confirmed patients in Chongqing, and 18 (24%) patients had no definite epidemiological link. The most frequent chronic medical illnesses were diabetes (27%) and hypertension (19%). The most common symptoms were cough (83%), fever (68%) and dyspnea (57%). 2 (3%) patients were under asymptomatic period before hospitalization, and appeared dyspnea without fever during the stay in hospital.

**Table 1.**
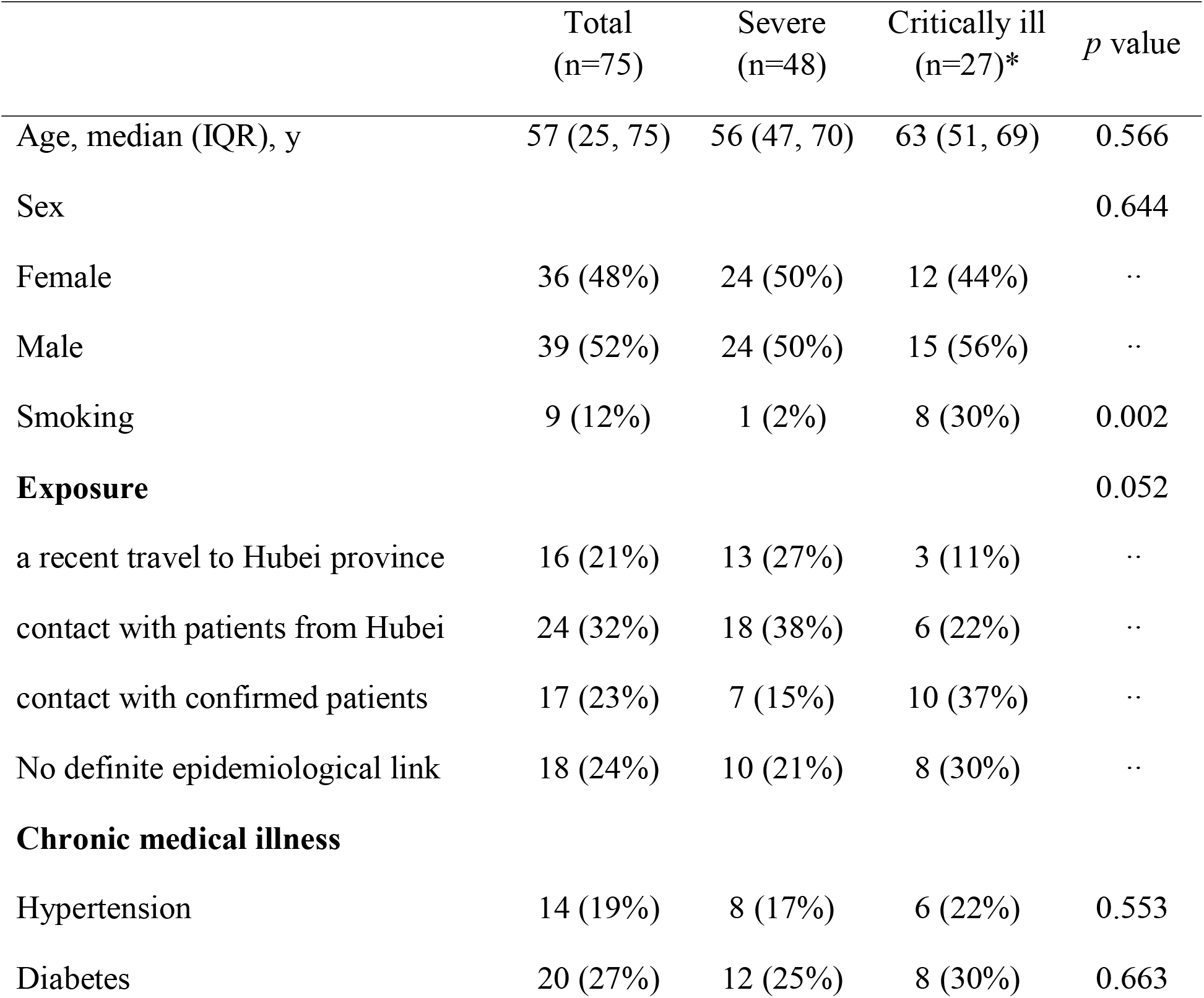
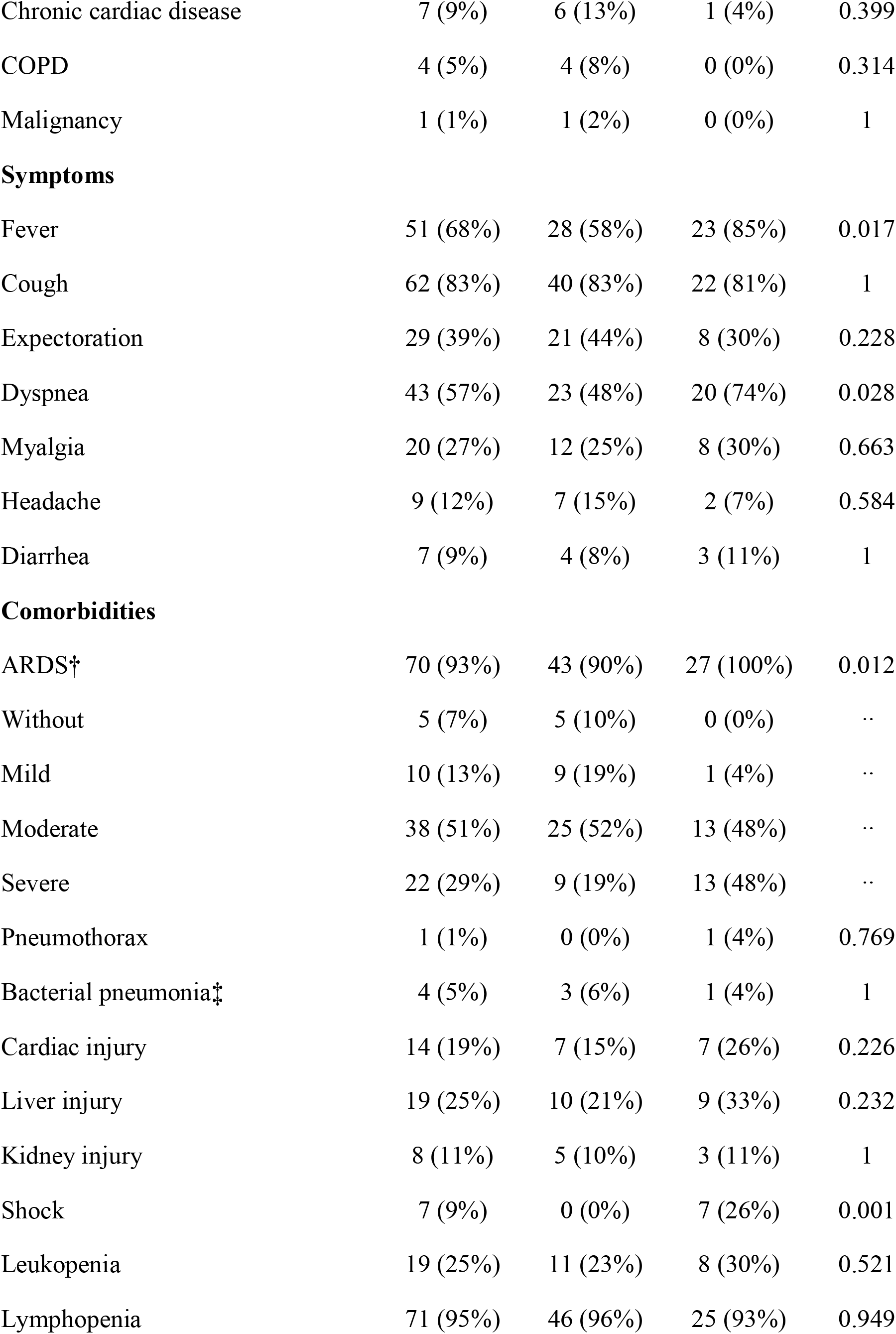
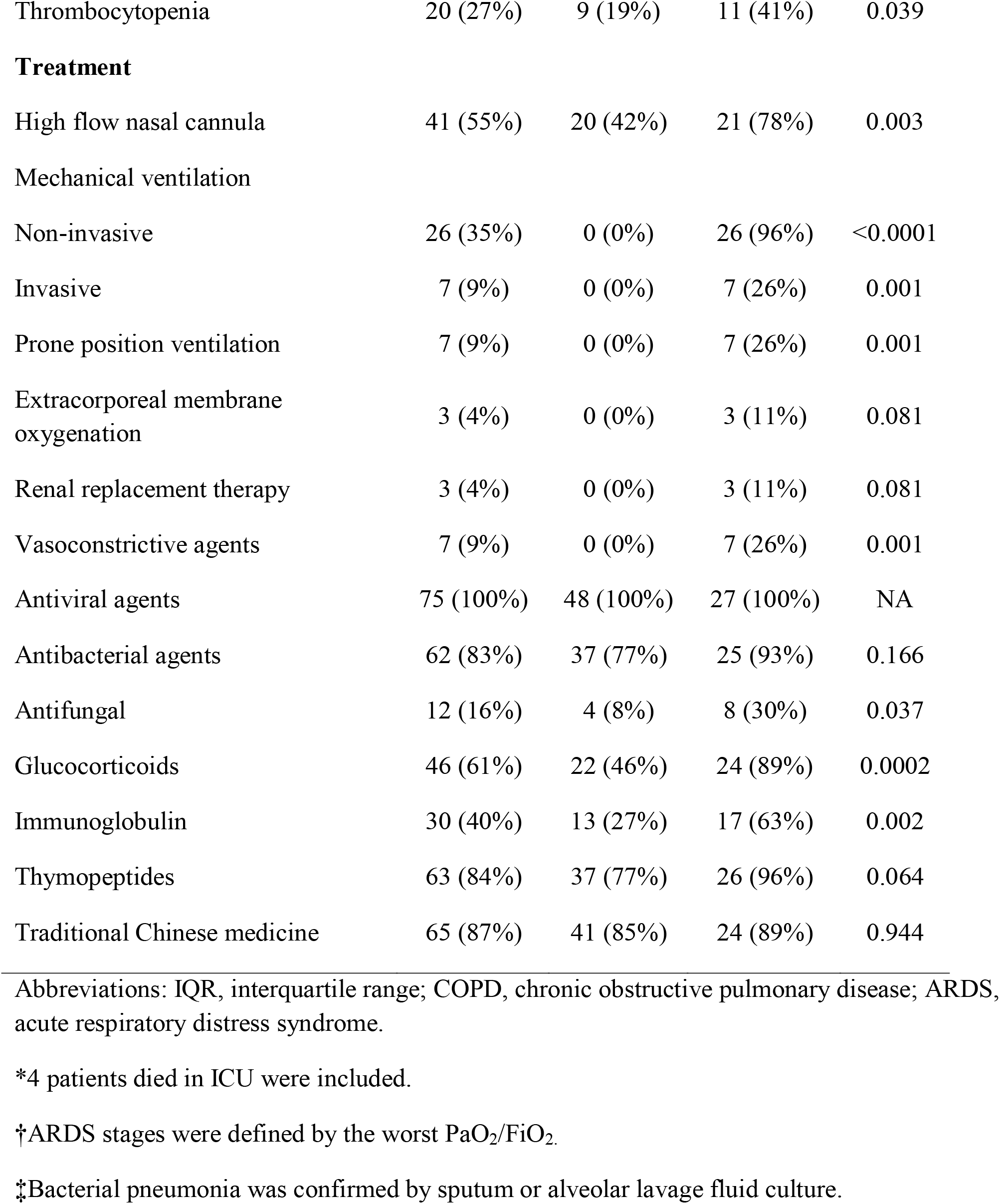
Clinical characteristics of ICU patients (severe/critically ill) with COVID-19.

ARDS was developed in most of the patients (93%), and more than half were moderate (Table 1). However, most of the patients (55%) supported with high flow nasal cannula (HFNC) oxygen therapy, 26 (35%) patients received non-invasive ventilation, and 7 (9%) patients received invasive ventilation. Other supportive treatments included: prone position ventilation in 7 (9%) patients, extracorporeal membrane oxygenation in 3 (4%), renal replacement therapy in 3 (4%), and vasoconstrictive agents in 7 (9%). Although bacterial pneumonia was identified by microbiological culture of sputum or alveolar lavage fluid in only 4 (5%) patients, the antibacterial agents were administered to 62 (83%) patients, and antifungal agents in 12 (16%) patients. Liver (25%) was the most commonly injured extrapulmonary organ, followed by cardiac (19%) and kidney (11%). Lymphopenia was a very noteworthy feature in these patients (95%), and lower incidence of leukopenia (25%) and thrombocytopenia (27%) relatively. Anti-viral agents were used in all patients (100%), the combination of Aluvia (Lopinavir and Ritonavir Tablets) and interferon alpha was the most commonly used. Traditional Chinese medicine was used in 65 (87%) patients owing to potential anti-viral and anti-inflammation activity. Glucocorticoids was given to 46 (61%) patients, immunoglobulin to 30 (40%), and hymopeptides to 63 (84%).

### Clinical course and outcomes

The clinical course and outcomes of patients with COVID-19 in Chongqing are shown in Table 2. Chongqing reported 6 deaths of COVID-19 up to 15 March 2020, with 1.04% mortality in all 576 patients. Because 2 patients were died in the emergency department, only 4 dead patients with clinical data were included in our study, with 5.3% 28-Day case fatality rate and 1.3% 28-Day mechanical ventilation dependency in ICU patients. The duration from any initial symptoms to diagnose confirmed by PCR test was 5 days (IQR 2-7), to hospital admission 7 days (IQR 4-8), to ARDS 7 days (IQR 6-10), to ICU admission 8 days (IQR 6-11), to ventilation 10 days (IQR 7-14), to viral shedding 20 days (IQR 16-26), and to death 16 days (min 15, max 28). The length of ICU stay was 13 days (IQR 9-19) and hospital stay 22 days (IQR 16-34).

**Table 2.** Clinical course and outcomes of ICU patients with COVID-19.

|  |  |
| --- | --- |
| Duration from any initial symptoms* | Median (IQR), days |
| to diagnose confirmed by PCR test | 5 (2, 7) |
| to hospital admission | 7 (4, 8) |
| to ARDS | 7 (6, 10) |
| to ICU admission | 8 (6, 11) |
| to ventilation | 10 (7, 14) |
| to viral shedding† | 20 (16, 26) |
| to death‡ | 16 (min 15, max 28) |
| Length of ICU stay | 13 (9, 19) |
| Length of hospital stay | 22 (16, 34) |
| Outcomes (n = 75) | NO. (%) |
| 28-Day Mortality‡ | 4 (5.3) |
| 28-Day mechanical ventilation dependency | 1 (1.3) |
| Location of death (n = 6) |  |
| ICU | 4 (66.7) |
| Emergency department | 2 (33.3) |

Abbreviations: PCR, polymerase chain reaction; ICU, intensive care unit; IQR, interquartile range.
\*2 patients without any symptoms until hospital admission were excluded for statistical analysis.
†The viral shedding was defined as two or more consecutive throat swab PCR test were negative.
‡4 patients died in ICU were included.

### Early improvement of ARDS

Learned from the clinical practice, we found a large group of ARDS patients would be improved in one week, we defined these patients as early improvement of ARDS (eiARDS). 56 patients who had ARDS (PaO2/FiO2 < 300mmHg) on the first day of ICU admission (Day 1) were included for analysis. We defined two groups based on the severity of illness on Day 7 (Fig. 1A): “eiARDS” patients PaO2/FiO2 ≥ 300mmHg, “Non-eiARDS” patients PaO2/FiO2 < 300mmHg. 18 patients got an eiARDS, accounted for nearly one third of the 56 ARDS patients. There are not significantly differences between the two groups in the PaO2/FiO2 on Day 1 (Fig. 1B), the proportion of pneumonia volume on Day 1(Fig. 1C), and the rate of ventilator usage (2 = 2.46, *p* = 0.117). Predictably and regrettably, all of the 4 dead patients did not get an early improvement of ARDS. What’s more, patients of “eiARDS” stayed shorter in ICU than the “Non-eiARDS”, with 10.5 days (IQR 8, 16) and 18 days (IQR 13, 22) respectively (*p* = 0.001) (Fig. 1D).

**Figure 1.**
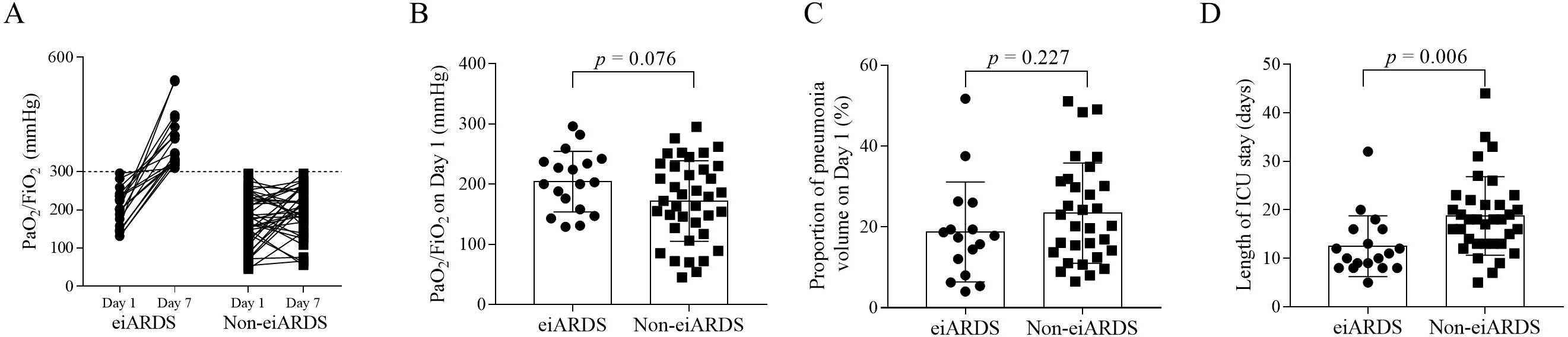
**Legend**. Comparison between “eiARDS” and “Non-eiARDS”. (A) All patients were under ARDS on Day 1, but divided into two groups (eiARDS and Non-eiARDS) according to the PaO2/FiO2 on Day 7. (B) No significantly difference between two groups in the PaO2/FiO2 on Day 1. (C) No significantly difference between two groups in the proportion of pneumonia volume on Day 1. (D) The length of ICU stay showed different between two groups. eiARDS, early improvement of ARDS; Non-eiARDS, none of eiARDS.

In order to determine the factors associated with eiARDS, we performed a bivariate analysis. As shown in Table 3, 3 variables (Age, Temperature and hsCRP) with p < 0.05 in the univariate analysis were chosen for multivariable analysis. Age (< 55 years) was the only variable independently associated with eiARDS, with an odds ratio of 7.4 (95%CI: 1.80-31.08). Indicating that patients younger than 55 years old are 7.4 times more likely to get an early improvement of ARDS than older ones.

**Table 3.** Univariate and multivariate analysis of predictors for early improvement of ARDS.

|  | Variable OR<br>(95% CI) | p value |
| --- | --- | --- |
| **Demographics and clinical characteristics** |  |  |
| Age, years* |  |  |
| ≥55 | 1 (ref) |  |
| <55 (univariate) | 4.33 (1.31-14.32) | 0.016 |
| <55 (multivariate) | 7.40 (1.80-31.08) | 0.006 |
| Male sex (vs female) | 2.00 (0.62-6.43) | 0.245 |
| Time of onset to ICU | 1.03 (0.87-1.22) | 0.704 |
| Smoke | 2.43 (0.53-11.10) | 0.252 |
| Hypertension | 0.55 (0.10-2.98) | 0.491 |
| Diabetes | 0.80 (0.21-3.01) | 0.741 |
| COPD | 0.69 (0.07-7.10) | 0.752 |
| APACHE II | 0.85 (0.70-1.05) | 0.130 |
| SOFA | 0.96 (0.56-1.67) | 0.889 |
| Temperature, °C* |  |  |
| <37.3 | 1 (ref) |  |
| ≥37.3 (univariate) | 0.22 (0.06-0.90) | 0.034 |
| ≥37.3 (multivariate) | 0.307 (0.06-1.68) | 0.173 |
| Heart rate, /min |  |  |
| <100 | 1 (ref) |  |
| ≥100 | 2.54 (0.63-10.25) | 0.191 |
| Respiratory rate, /min |  |  |
| <24 | 1 (ref) |  |
| >=24 | 1.27 (0.32-5.04) | 0.738 |
| Systolic pressure, mmHg |  |  |
| <140 | 1 (ref) |  |
| >=140 | 1.21 (0.47-3.10) | 0.695 |
| Ventilation (vs Non-) | 0.83 (0.14-4.73) | 0.829 |
| Proportion of pneumonia volume, % | 0.97 (0.92-1.02) | 0.227 |
| **Laboratory findings** |  |  |
| pH |  |  |
| 7.35-7.45 | 1 (ref) |  |
| >7.45 | 1.68 (0.46-6.21) | 0.437 |
| PaCO<sub>2</sub>, mmHg |  |  |
| 34-45 | 1 (ref) |  |
| <34 | 1.10 (0.35-3.46) | 0.873 |
| PaO<sub>2</sub>, mmHg |  |  |
| >=60 | 1 (ref) |  |
| <60 | 0.44 (0.12-1.59) | 0.209 |
| White blood cell count, × 10<sup>3</sup>/L |  |  |
| <4 | 3.63 (0.97-13.64) | 0.056 |
| 4-10 | 1 (ref) |  |
| >10 | 2.07 (0.30-14.44) | 0.461 |
| Lymphocyte, ×10<sup>3</sup> | 2.73 (0.54-13.80) | 0.224 |
| Platelet, × 10<sup>3</sup>/L |  |  |
| >=100 | 1 (ref) |  |
| <100 | 2.19 (0.28-16.95) | 0.454 |
| Potassium, mmol/L |  |  |
| 3.5-4.5 | 1 (ref) |  |
| <3.5 | 0.51 (0.12-2.12) | 0.352 |
| Sodium, mmol/L |  |  |
| 135-145 | 1 (ref) |  |
| <135 | 1.05 (0.34-3.27) | 0.933 |
| Creatinine, $\mu\text{mol/L}$ | 1.00 (0.97-1.03) | 0.804 |
| Albumin, g/L |  |  |
| $\geq 40$ | 1 (ref) | |
| <40 | 0.57 (0.11-2.87) | 0.491 |
| Total bilirubin, $\mu\text{mol/L}$ | | |
| $\leq 17.1$ | 1 (ref) | |
| >17.1 | 1.96 (0.61-6.29) | 0.256 |
| Alanine aminotransferase, U/L |  |  |
| $\leq 40$ | 1 (ref) | |
| >40 | 0.59 (0.17-2.00) | 0.396 |
| Creatine kinase, U/L |  |  |
| $\leq 200$ | 1 (ref) | |
| >200 | 0.72 (0.17-3.10) | 0.657 |
| High-sensitivity cardiac troponin T, ng/mL, ng/mL |  |  |
| $\leq 0.014$ | 1 (ref) | |
| >0.014 | 0.70 (0.06-7.74) | 0.774 |
| Prothrombin time, sec |  |  |
| $\leq 14$ | 1 (ref) | |
| >14 | 2.53 (1.23-5.19) | 0.999 |
| D-dimer, µg/L |  |  |
| ≤0.55 | 1 (ref) |  |
| >0.55 | 0.94 (0.27-3.26) | 0.928 |
| Procalcitonin, ng/ml |  |  |
| ≤0.046 | 1 (ref) |  |
| >0.046 | 0.32 (0.09-1.17) | 0.084 |
| hsCRP, mg/L* (univariate) | 0.99 (0.98-1.00) | 0.041 |
| hsCRP, mg/L (multivariate) | 0.99 (0.98-1.00) | 0.092 |

Abbreviations: OR, odds ratio; ICU, intensive care unit; COPD, chronic obstructive pulmonary disease; APACHE, acute physiology and chronic health evaluation; SOFA, sequential organ failure assessment; PaCO<sub>2</sub>, arterial partial pressure of carbon dioxide; PaO<sub>2</sub>, arterial partial pressure of oxygen; hsCRP, high-sensitivity C reaction protein.
\* 3 variables (Age, Temperature and hsCRP) were chosen for multivariable analysis.

## Discussion

In the present study, we found that the mortality of COVID-19 in Chongqing was 1.04%, and the 28-Day case fatality rate of ICU patients was 5.3%. ARDS was developed in 93% ICU patients, and the HFNC was the most commonly used oxygen therapy. An early improvement of ARDS was occurred in about one third patients, and patients younger than 55 years old would be more likely to do this.

Mortality of COVID-19 varies widely in different periods and areas. In the early stage of outbreak, Wuhan reported 4.3% mortality in hospitalized patients(14), and 61.5% in critically ill patients(15). However, the mortality of ICU patients gradually decreased to 32.5-38.5% as time elapsed(16,17). This happened to be 26% in Lombardy Region, Italy(18) and 50% in Seattle Region, America(19). In the present study, we found that only 5.3% case fatality rate of ICU patients was occurred in Chongqing. The big differences of the mortality probably due to whether the medical resources can be timely supplied, including heath workers and hospital beds(20). As a matter of fact, a huge number of health workers from other provinces have been aided to Hubei province, with consecutively increased acute care beds(20). Similar to the model of Hubei, as the first cluster cases of COVID-19 was detected in Chongqing, 4 designated hospitals were arranged and prepared for patients with COVID-19 only, and also the medical experts from different hospitals in Chongqing. The centralized dispatcher of medical resources is the key experience of treating COVID-19 in China.

ARDS is the key factor to affect the mortality. According to the Berlin definition, stages of mild, moderate, and severe ARDS were associated with increased mortality (27%, 32% and 45% respectively)(11). Studies have shown that ARDS was one of the risk factors of death in patients with COVID-19(15,21). Many efforts have been attempted to treat ARDS. However, only mechanical ventilation was showed to be effective therapeutics(22). Interestingly, in our present study, although 93% patients had ever suffered ARDS, only 35% patients received non-invasive ventilation, and 9% patients received invasive ventilation. The most commonly used oxygen therapy was HFNC, which counted for 55% (despite patients may receive both HFNC and ventilation), seemly meaning that HFNC is effective for COVID-19-induced ARDS. Similar conclusions can be observed in a previous review(4). The authors hold that HFNC is suitable for COVID-19 patients with mild ARDS, and even safe for moderate and severe patients, which is clearly inconsistently with the stratified treatment strategies of ARDS caused by other factors(4).

In our present study, we found nearly one third patients with ARDS recovered in one week, which we defined as an early improvement of ARDS (eiARDS). However, this eiARDS can be found in only 18% patients with mild ARDS caused by other factors, with 36% patients persisting and 46% worsened in the first week after ARDS onset(23). It is worth mentioning that, why so many patients with COVID-19 got an eiARDS and why HFNC oxygen therapy was so effectual for these patients? There is a voice should be considered. Gattinoni et al.(24) proposed two types of patients with COVID-19 pneumonia: “non-ARDS,” type 1, and ARDS, type 2. Although both types of patients meet the ARDS Berlin definition, but the severe hypoxemia in type 1 patients is associated with nearly normal respiratory system compliance, which can lead to ventilation/perfusion mismatch(24). Assuming that type 1 pneumonia would improve quicker than the type 2, it appears to be able to explain the above problems. In addition, Gattinoni et al proposed that the gas volume and percentage of non-aerated tissue can be clearly distinguished by CT scan between type 1 and type 2 pneumonia. However, there was not any differences in the proportion of pneumonia volume between “eiARDS” and “Non-eiARDS” in our study.

Regardless of the reasons why the proportion of eiARDS was so high, paying attention to eiARDS itself is clinically meaningful. Early or rapidly improving ARDS always associated with a better surviving or outcomes(23,25). For COVID-19 patients, early improvement in oxygenation was associated with being discharged alive from the ICU(21). We found that patients with eiARDS had a higher survival rate and lower length of ICU stay than the “Non-eiARDS”. Dynamic observation of ARDS in the short term is very worthwhile for the prognosis of COVID-19, and patients whose ARDS did not improve in one week should be given more attention.

One might point out that whether it is because most patients had mild ARDS at baseline, so it is easier to reach eiARDS. But the fact is that, in the baseline, 38.8% patients were under moderate ARDS, and no significantly differences were shown between “eiARDS” and “Non-eiARDS. In other words, the initial PaO2/FiO2 was independently associated with eiARDS. Indeed, multiple studies have shown that older age (> 65 years) is one of the risk factors of death in patients with COVID-19(15,26–28), and establishing risk stratification through age (> 60 years) maybe helpful to clinicians(29). Similar underlying mechanisms may be found in regard to the effect of age to death and the development of ARDS. Nevertheless, age should be given high attention during the management of COVID-19 patients.

## Limitations

This study has several limitations. First, based on the retrospective study design, the laboratory tests (except arterial blood gas analysis, which performed each day) may not be performed in all patients at a specific time, and the missing data was replaced by the values within latest 2 days. Second, although the treatment strategies of the two hospitals followed the guidelines issued by the Chinese National Health Commission, some of the treatments are different, such as the composition of traditional Chinese medicine, which may cause different clinical outcomes. Third, the sample size is relatively small, part of the conclusions needs to be verified by multiple centers and larger sample size.

## Conclusions

In the present study, we described the epidemiology, clinical features, laboratory data, treatments, and outcomes of ICU patients in Chongqing, China. We found that the case fatality rate of ICU patients in this region was only 5.3%, and the timely supplement of medical resources and oxygen therapy based on HFNC may be the reasons for this low fatality rate. In addition, we identified a new subphenotype of ARDS – the early (in one week) improvement of ARDS (eiARDS), which occurred in about one third of ARDS patients with COVID-19, anticipating a favorable clinical outcome. The age (< 55 years) is an independent predictor for the eiARDS, and stratification of COVID-19 patients by age is recommended.

## Data Availability

The datasets generated and/or analyzed during the current study are not publicly available due to additional papers that will be published on other related topics but are available from the corresponding author on reasonable request.

## Competing interests

The authors declare that they have no competing interests.

## Funding

This work was supported by the Emergency Research Project of COVID-19 of Chongqing Health Commission (2020NCPZX04).

## Acknowledgments

We thank the patients described in this report, the health care personnel who cared for them, the staff members of Health Commission of Chongqing City, and the staff members of Chongqing Center for Disease Control and Prevention.

## Notes

### Competing Interest Statement

The authors have declared no competing interest.

### Author Declarations

The study was approved by the Research Ethics Commission of the second affiliated hospital of Chongqing medical university, Chongqing public health medical center and Chongqing three gorges central hospital.

